# Cost-effectiveness of internet-delivered cognitive behaviour therapy for body dysmorphic disorder: results from a randomised controlled trial

**DOI:** 10.1101/2021.08.23.21262455

**Authors:** Oskar Flygare, Erik Andersson, Gjermund Glimsdal, David Mataix-Cols, Diana Djurfeldt, Christian Rück, Jesper Enander

## Abstract

**Objectives:** To evaluate the cost-effectiveness of internet-delivered cognitive behaviour therapy for body dysmorphic disorder (BDD-NET).

**Design:** Secondary cost-effectiveness analysis from a randomised controlled trial on BDD-NET versus online supportive psychotherapy.

**Setting:** Academic medical centre.

**Participants:** Self-referred adult patients with a primary diagnosis of body dysmorphic disorder and a score of 20 or higher on the modified Yale-Brown obsessive compulsive scale (n = 94). Patients receiving concurrent psychotropic drug treatment were included if the dose had been stable for at least two months and remained unchanged during the trial.

**Interventions:** Participants received either BDD-NET (n = 47) or online supportive psychotherapy (n = 47) for 12 weeks.

**Primary and secondary outcome measures:** The primary outcome measures were cost-effectiveness and cost-utility from a societal perspective, using remission status from a diagnostic interview and quality-adjusted life years from EQ-5D, respectively. Secondary outcome measures were cost-effectiveness and cost-utility from a health care perspective and the clinic’s perspective.

**Results:** Compared to supportive psychotherapy, BDD-NET produced one additional remission for an average societal cost of $4132. The cost-utility analysis showed that BDD-NET generated one additional QALY to an average cost of $14319 from a societal perspective.

**Conclusions:** BDD-NET is a cost-effective treatment for body dysmorphic disorder, compared to online supportive psychotherapy. The efficacy and cost-effectiveness of BDD-NET should be directly compared to face-to-face cognitive behaviour therapy.

**Trial registration:** NCT02010619

**Strengths and limitations of this study:**

- This is the first cost-effectiveness evaluation of a novel internet-delivered treatment designed to increase access to cognitive behaviour therapy for patients with body dysmorphic disorder
- Cost estimations were self-rated and might not capture all costs associated with treatment
- The self-referred participants were likely motivated to undergo treatment.

## Background

Body dysmorphic disorder (BDD) is characterised by an intense pre-occupation with perceived flaws in appearance that are not noticeable or appear slight to others. Cognitive behaviour therapy (CBT) is an effective treatment for BDD (1– 4), but few patients receive this evidence-based treatment for BDD. In one online survey, only 18% of patients with BDD had ever received CBT (5). Barriers to treatment include shame and stigma associated with BDD, the costs of treatment, and belief that a treatment would not work (5,6). In addition, there is a shortage of specialised CBT therapists, particularly in areas far away from universities and CBT training centres (7). Thus, specialist CBT for BDD is inaccessible for most patients.

Internet-delivered cognitive behaviour therapy (ICBT) has been developed for a range of mental health disorders in an effort to improve access to evidence-based treatment (8). ICBT interventions include the same content as regular CBT and differ only in the mode of delivery: the patient logs into a secure web-based platform to access treatment content and communicate with a therapist. Previous studies have shown that therapist-guided ICBT for BDD (BDD-NET) is an efficacious treatment for adults with BDD, with effects sustained up to two years after treatment (9–11). An advantage of remote treatments such as BDD-NET is that they require less therapist time per patient, enabling more patients to access therapy and reducing costs. However, the cost-effectiveness of BDD-NET has not been formally evaluated. Here, we analysed cost data from our controlled trial (9) and evaluated the cost-effectiveness of BDD-NET compared to online supportive psychotherapy. We hypothesised that BDD-NET would be cost-effective from societal, health care, and the clinic’s perspectives.

## Methods

### Trial design

This study reports planned secondary cost-effectiveness analyses from a randomised controlled trial comparing BDD-NET to supportive psychotherapy (N = 94) (9). The study was approved by the regional ethical review board in Stockholm (2013/1773-31/4) and pre-registered at clinicaltrials.gov (NCT02010619). Recruitment, treatment, and follow-up in the original study was conducted between November 2013 and January 2015. The results are reported in accordance with the CHEERS reporting guidelines for health economic evaluations (12).

### Participants

Included participants had a primary diagnosis of BDD according to the Diagnostic and Statistical Manual of Mental Disorders, 5th edition (DSM-5; (13)), were at least 18 years old, had access to the internet, and a score of 20 or above on the modified Yale-Brown obsessive-compulsive scale for BDD (BDD-YBOCS; (14)). Participants who were taking psychotropic drugs were asked to keep their dose stable during the study period. Exclusion criteria were initiation or changes in psychotropic drug treatment within two months before enrolment, completed CBT for body dysmorphic disorder within the past 12 months, ongoing psychological treatment, current substance dependence, bipolar disorder or psychosis, acute suicidal ideation, or a severe personality disorder that could jeopardize participation in treatment (such as borderline personality disorder with self harm).

### Interventions

#### BDD-NET

BDD-NET was delivered through a secure online platform hosted at a dedicated hospital server with encrypted traffic. The treatment was based on a validated CBT protocol for body dysmorphic disorder (15), contained eight text-based modules accompanied by worksheets and homework assignments, and lasted 12 weeks. Participants were asynchronously supported by a therapist through a built-in email system on the BDD-NET webpage and could log in to send emails at any time. Therapists would then review and respond to messages within 36 hours on weekdays.

The therapists involved were four clinical psychology students with basic clinical training but with no prior experience of treating BDD. They received weekly supervision sessions with an experienced clinician (author JE) who also monitored messages on the platform to ensure treatment integrity and adherence to protocol.

#### Online supportive psychotherapy

Participants receiving online supportive psychotherapy had access to the email system on the BDD-NET webpage and received support in a similar way to BDD-NET. The therapists used counselling techniques such as reflecting, empathising and summarising in order to provide the same level of caregiver attention as BDD-NET. Notably, the supportive therapy did not include core parts of BDD-NET such as exposure with response prevention. For more detailed descriptions of the interventions, please see the main outcome paper (9).

### Measures

#### Clinical outcomes

The cost-effectiveness analyses were based on remission status and quality-adjusted life years (QALYs). Remission was defined as no longer meeting diagnostic criteria for body dysmorphic disorder in a structured interview with a clinician, blinded to treatment allocation. QALYs were estimated based on the self-rated EuroQol (EQ-5D) which is divided into five health domains: mobility, self-care, pain/discomfort, daily activities, and anxiety/depression. An index QALY score is then calculated ranging from 0 (dead) to 1 (perfect health) (16).

#### Health economic measures

We used the Trimbos questionnaire for costs associated with psychiatric illness (TIC-P) to gather information on self-rated costs in the past month (17). Costs were analysed from a societal perspective by including all direct and indirect, medical and non-medical costs (e.g., productivity losses, sick leave). From a health care perspective, costs directly associated with treatment and costs from other health care visits and medications were included. Finally, costs from the clinic’s perspective included only the cost of providing treatment (e.g., therapist salary). Costs for medications were calculated from market prices, and health care visit costs were based on tariffs from official listings for the Swedish health care system (see supplemental materials). Productivity losses were based on the mean gross earning in Sweden in 2014 (188 SEK), and domestic work cutback was estimated using the hourly tariff of €14 (18). The TIC-P was administered at pre-treatment, post-treatment and follow-up. Costs were summarised in Swedish Crowns, extrapolated to 12 weeks in order to cover the full duration between assessments, and converted to US dollars based on purchasing power parities for the year 2014 where 6.861 SEK paid one USD.

### Statistical analyses

All analyses were intention-to-treat with the assumption that data were missing at random. Costs from the three perspectives (societal, health care, clinic) were analysed with remission status and QALY change from pre-treatment to post-treatment and follow-up as outcomes, respectively. Mixed-effects linear regression for repeated measures were used for outcomes and costs, with fixed effects of group and time, their interaction, as well as a random intercept term. Missing values were estimated using maximum likelihood estimation in the mixed-effects models, and non-parametric bootstrapping (1000 replications) was used to estimate group differences which were based on the group * time interaction effects. The “net benefit” approach was used by estimating the probability of cost-effectiveness under different societal willingness-to-pay scenarios (19). Standardised Cohen’s *d* effect sizes were estimated for both within-group and between-group changes based on the least-squares means from the models, using the residual standard deviation of random effects as sigma (20). We used R version 4.0.2 (21) for all analyses and the analytical code is available on the Open Science Framework https://doi.org/10.17605/OSF.IO/DC69E.

## Results

### Completion rates and participant characteristics

A majority of the participants were female (n = 80 (85%)) and the average age of all the participants was 32.6 (SD = 11.97). See table 1 for demographic and baseline assessment information for both treatment groups. All 94 participants completed the post-treatment diagnostic interview, and at follow-up, 85 (90%) participants completed the diagnostic interview (BDD-NET: 40/47 = 85%, supportive therapy: 45/47 = 96%). Completion rates for EQ-5D were similar, with 92 (98%) assessments (BDD-NET: 45/47 = 96%, supportive therapy: 47/47 = 100%) at post treatment, and 80 (85%) assessments completed at follow-up (BDD-NET: 36/47 = 77%, supportive therapy: 44/47 = 94%). Completion rates for the TIC-P cost assessment were high with 91 (97%) responses at post treatment (BDD-NET: 45/47 = 96%, supportive therapy: 46/47 = 98%) and 79 (84%) responses at follow-up (BDD-NET: 36/47 = 77%, supportive therapy: 43/47 = 91%). Figure 1 shows participant flow through the different assessment points.

**Table 1.** Sample characteristics

| Table 1. Sample characteristics |  |  |
| --- | --- | --- |
|  | Online supportive therapy | BDD-NET |
| n | 47 | 47 |
| Female, n (%) | 41 (87%) | 39 (83%) |
| Age (mean (SD)) | 31.32 (10.12) | 33.87 (13.55) |
| Education, n (%) |  |  |
| Primary school | 6 (13%) | 4 (9%) |
| High school | 23 (49%) | 31 (66%) |
| College/university | 17 (36%) | 11 (23%) |
| Doctorate | 1 (2%) | 1 (2%) |
| SSRI, n (%) | 8 (17%) | 5 (11%) |
| Pre-treatment BDD-YBOCS (mean (SD)) | 28.51 (4.55) | 29.13 (5.02) |
| Pre-treatment EQ-5D (mean (SD)) | 0.75 (0.18) | 0.71 (0.22) |

**Figure 1.**
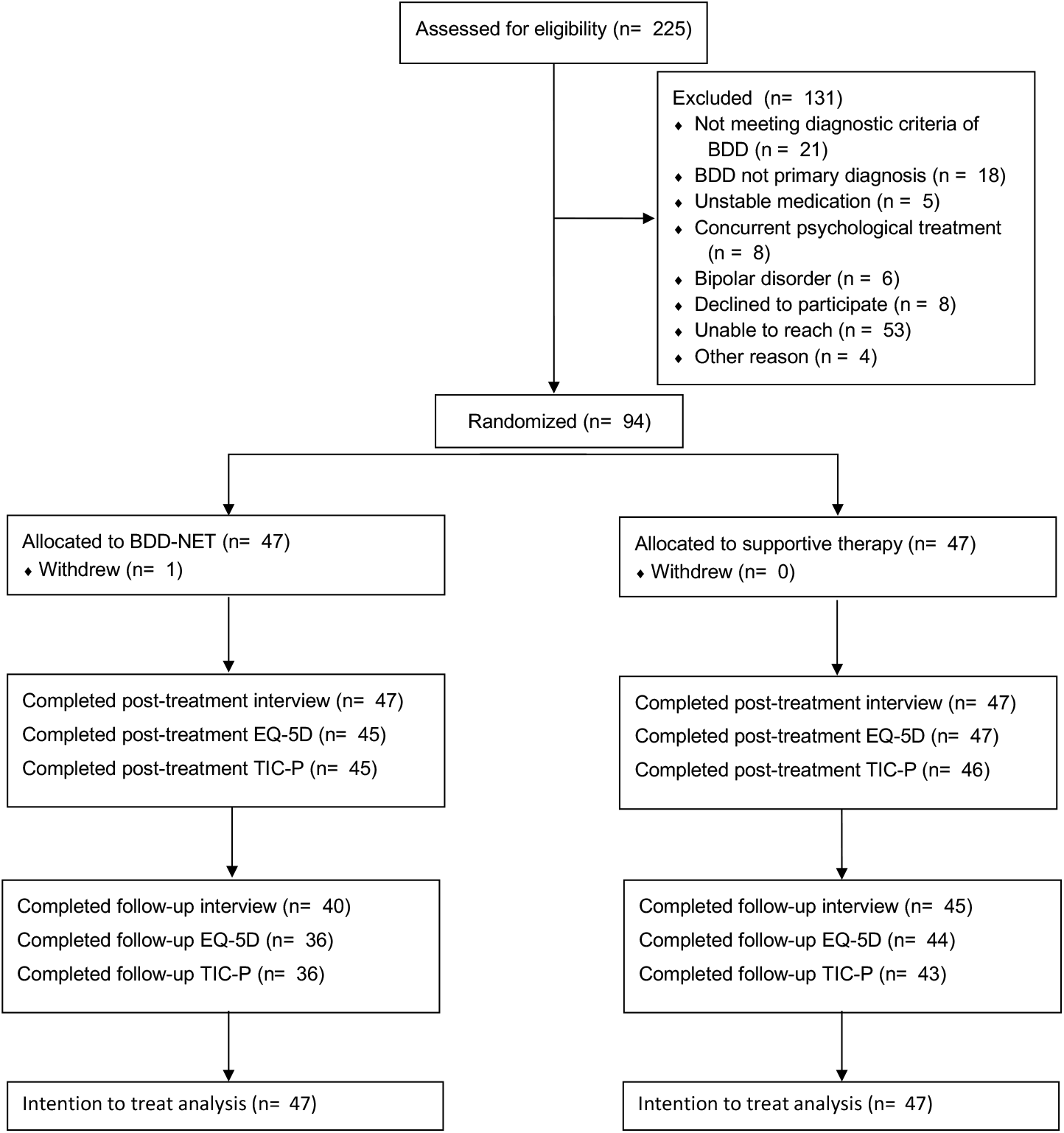
Diagram of participant flow and completion of measures. Abbreviations: BDD-NET, internet-delivered therapist-guided cognitive behaviour therapy for body dysmorphic disorder; EQ-5D, EuroQol 5-Dimensions; TIC-P, Trimbos questionnaire for costs associated with psychiatric illness.

### Clinical efficacy

The proportion of remitters at post-treatment, i.e. participants that no longer met diagnostic criteria according to the structured interview, were 32% [95% CI 20 to 46] in the BDD-NET group and 2% [95% CI 0 to 14] in the group that received supportive therapy. Patients in the BDD-NET group were more likely to be in remission at post-treatment (OR = 3.07, SE = 1.06, *p* = .004). At follow-up, results were similar with a higher proportion of remitters in the BDD-NET group 40% [95% CI 26 to 56] compared to supportive therapy 9% [95% CI 3 to 21] and corresponding difference in a direct comparison (OR = 1.92, SE = 0.62, *p* = .002).

EQ-5D QALY estimates at post-treatment were on average 0.73 [95% CI 0.66 to 0.8] in the BDD-NET group and 0.67 [95% CI 0.61 to 0.74] in the supportive therapy group, with a small increase from pre-treatment in the BDD-NET group (*d* = 0.08 [95% CI -0.34 to 0.5]) and a medium-sized decrease in the supportive therapy group (*d* = -0.39 [95% CI -0.8 to 0.03]). At follow-up, the estimates were largely unchanged with an average of 0.8 [95% CI 0.72 to 0.87] in the BDD-NET group and 0.67 [95% CI 0.6 to 0.74] in the group that received supportive therapy. Within-group effect sizes from pre-treatment to follow-up were *d* = 0.46 [95% CI 0 to 0.91] for BDD-NET and *d* = -0.41 [95% CI -0.83 to 0.01] for supportive therapy. The between-group effect sizes were small at post-treatment (*d* = 0.29 [95% CI -0.22 to 0.8]) and medium at follow-up (*d* = 0.69 [95% CI 0.15 to 1.24]).

### Costs

Costs from the societal perspective (e.g., including all direct, indirect, medical and non-medical costs) increased in the BDD-NET group (*b* = $928) and decreased in the supportive therapy group (*b* = $-302) groups over the course of treatment, and decreased in both groups between post-treatment and follow-up (BDD-NET: *b* = $-386, supportive therapy: *b* = $-243). Similarly, when looking at costs from a health care perspective (e.g., direct medical costs from visits and medications), costs from pre-to post-treatment increased in BDD-NET (*b* = $982) and were stable in supportive therapy (*b* = $-55). From post-treatment to follow-up, costs were stable (BDD-NET: *b* = $103) or decreased slightly (supportive therapy: *b* = $-269). Detailed descriptions of costs for all sub-categories are shown in the supplemental materials.

The cost of providing treatment from the clinic’s perspective included the salary cost of therapists and was estimated to $946 for BDD-NET and $461 for supportive therapy. This cost corresponds to an average time of 15.7 minutes per patient and week for BDD-NET and 7.7 minutes for supportive therapy.

### Cost-effectiveness and cost-utility

#### The societal perspective

The cost-effectiveness of BDD-NET versus the supportive therapy control treatment was investigated up until the 3-month controlled follow-up, after which the control group was offered BDD-NET. The cost-effectiveness ICER at post-treatment was 1231 / 0.3 = $4132 per case in remission. At follow-up, the corresponding numbers were 1088 / 0.29 = $3731 per case in remission. BDD-NET was cost-effective from a societal perspective, given that society is willing to pay up to $4132 per case in remission after treatment. A majority of ICERs from bootstrapped samples were located in the northeast quadrant, confirming higher efficacy and costs of BDD-NET compared to online supportive therapy (Figure 2A).

**Figure 2.**
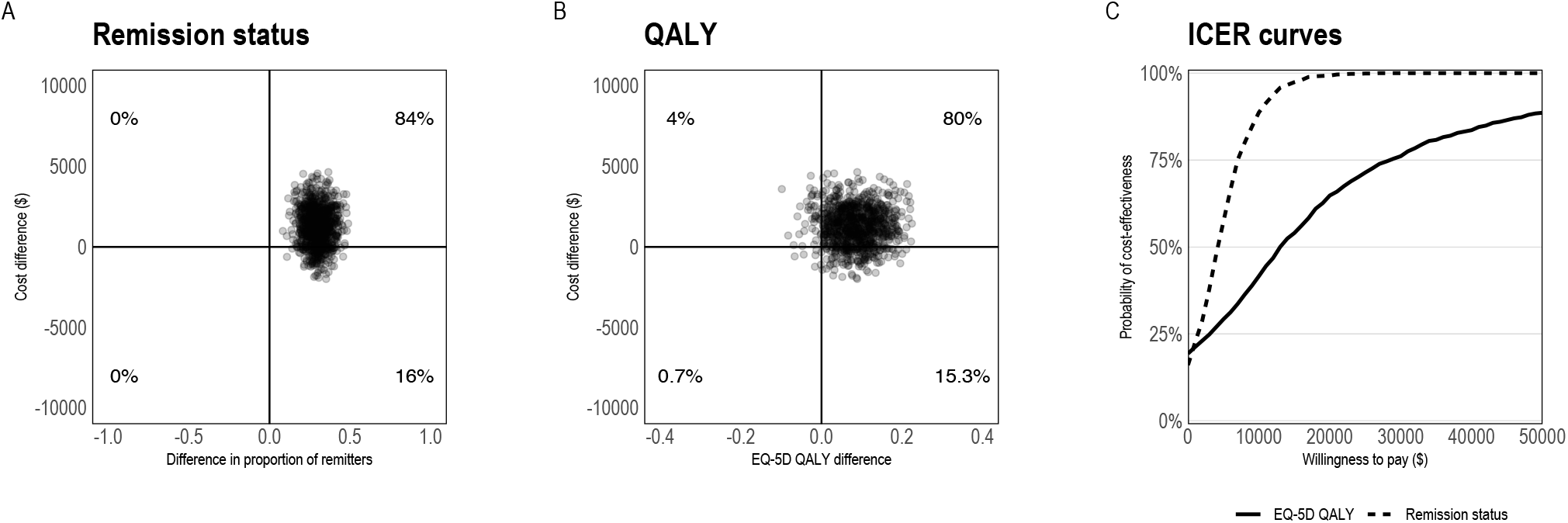
Pre-treatment to post-treatment cost-effectiveness of BDD-NET compared to supportive therapy. Cost-effectiveness planes for BDD remission rates (2A), QALYs (2B), and acceptability curves (2C). Costs are from a societal perspective and based on the TIC-P questionnaire using Swedish health care tariff listings and national statistics. Abbreviations: BDD-NET, internet-delivered cognitive behaviour therapy for body dysmorphic disorder; ICER, incremental cost-effectiveness ratio; QALYs, Quality-adjusted life years.

Cost-utility analyses (analyses of QALYs in relation to costs) indicated that the societal cost for one additional QALY gain from BDD-NET versus supportive therapy was 1231 / 0.09 = $14319 at post-treatment, and 1088 / 0.16 = $6784 at follow-up. Given the small differences in EQ-5D QALY gain, BDD-NET achieved 75% cost-utility at a societal willingness to pay of $29000 at post-treatment (Figure 2C). Again, the majority of ICERs from bootstrapped samples were located in the northeast quadrant of the cost-effectiveness plane (Figure 2B).

#### The health care perspective

From the health care perspective, which includes costs of health care and medications (but excludes non-medical or indirect costs), the cost-effectiveness of BDD-NET at post treatment was 1037 / 0.3 = $3482 per case in remission. The cost-effectiveness of BDD-NET versus supportive therapy at follow-up was 1409 / 0.29 = $4831. Compared to supportive therapy, BDD-NET was estimated to have an additional cost of $3482 per case in remission, and the ICERs from bootstrapped samples were located in the northeast quadrant of the cost-effectiveness plane (see supplemental materials).

#### The clinic’s perspective

The clinic’s perspective includes the cost of delivering the interventions (in this case, the cost of the psychologists’ time). The cost-effectiveness of BDD-NET compared to supportive therapy at post-treatment was 485 / 0.3 = $1628 per case in remission. At follow-up, cost-effectiveness was 485 / 0.29 = $1662. Again, the ICERs from bootstrapped samples were located inn the northeast quadrant of the cost-effectiveness plane.

## Discussion

To our knowledge, this is the first cost-effectiveness study of a psychological treatment for BDD. We found that therapist-guided internet-delivered cognitive behaviour therapy for body dysmorphic disorder (BDD-NET) produced one additional remission for an average societal cost of $4132 and the cost-utility analysis showed that BDD-NET generated one additional QALY to an average price of $14319 from a societal perspective, compared to online supportive therapy. Results were similar when costs from a health care perspective and the clinic’s perspective were considered.

Previous studies on the cost-effectiveness internet-delivered cognitive behaviour therapy (ICBT) for related disorders have found similar results. For example, (22) found that an internet-delivered CBT for obsessive-compulsive disorder produced an additional remission for $672 and one additional QALY for $4800 compared to online supportive therapy. In cost-effectiveness analyses of ICBT for health anxiety, savings of £1244 per additional case of remission were found compared to an attention control condition (23), and several forms of low-intensity treatments were found to be cost-effective compared to waiting list (£-134 to £416 per additional case in remission) (24). Similarly, ICBT for depression was associated with a cost of €1817 for each additional reliably improved participant (25) and has been estimated to be cost-effective at a societal willingness to pay of £20 000 per additional QALY compared to usual care (26). Caution should be taken in directly comparing results from different studies since they may differ in the estimation of costs, outcome definitions, health care context, or comparators. Keeping this limitation in mind, a meta-analysis of ICBT for various conditions found that the treatments are cost-effective options by providing large clinical gains at relatively low cost (27).

The results should be viewed in light of several limitations. First, the current study used self-reported costs from the TIC-P, which might not capture all costs associated with treatment. However, the cost estimations from the TIC-P are reliable and correspond well to costs estimated from medical registrations (28). Second, BDD-NET was compared to supportive psychotherapy which is not an established treatment for BDD, however face to face supportive psychotherapy was associated with comparable gains to CBT for BDD in a recent randomised controlled trial, indicating that it is an effective treatment for some patients with BDD (3). Third, participants in this study were self-referred and thus likely to be motivated to undergo treatment. This may not be representative for the whole population of patients with BDD, who may have poor insight and be less motivated to seek help for their problems.

Future research should directly compare the efficacy and cost-effectiveness of BDD-NET to face-to-face CBT for BDD, which together with SSRI medication, is a first line treatment for BDD. Additionally, evaluation of BDD-NET in regular psychiatric care is needed as these patients may differ in level of insight, psychiatric comorbidity and willingness to undergo psychological treatment for their BDD, compared to self-referred participants.

In conclusion, BDD-NET is a cost-effective treatment for BDD, compared to online supportive psychotherapy. The results have direct implications for rational decision-making in health care, where information on the cost-effectiveness of treatment alternatives can improve the distribution of limited resources, thus providing effective treatment to more patients at a minimal cost.

## Supporting information

CHEERS checklist

## Data Availability

The datasets analysed are not publicly available since they contain sensitive personal identifying information and data sharing was not part of the written informed consent, but are available from the corresponding author on reasonable request. Statistical code is publicly available from the Open Science Framework repository: https://doi.org/10.17605/OSF.IO/DC69E

## Article information

### Ethical approval

The regional ethical review board in Stockholm approved the protocol (registration ID: 2013/1773-31/4). All participants gave written informed consent.

### Contributors

JE and CR had the original idea for the study and, with DM-C, designed the trial variables and obtained funding. JE and CR were responsible for study supervision. OF carried out the statistical analysis and drafted the manuscript, which was revised by EA, GG, DM-C, DD, CR and JE. All authors approved the final version to be submitted.

