## Supplemental materials for "Cost-effectiveness of internet-delivered cognitive behaviour therapy for body dysmorphic disorder: results from a randomised controlled trial"

### Supplemental methods

#### Cost sources

Costs for medications were estimated based on the prices in The Dental and Pharmaceutical Benefits Agency database, which is publicly available online (<https://www.tlv.se/beslut/sok-i-databasen.html>). If dose or product name was not reported in TIC-P (for example responding “SSRI”), costs were estimated based on the standard dose of the most common product. Costs for alternative medications were based on market prices. Standard costs for health care visits are shown in table A.1.

| Table A.1. Standard costs for health care visits | | |
| --- | --- | --- |
| Health service | Standard cost | Unit |
| General practitioner | $ 320 | Consultation |
| Company physician | $ 175 | Consultation |
| Psychiatrist | $ 662 | Consultation |
| Medical specialist (other) | $ 446 | Consultation |
| District nurse | $ 107 | Consultation |
| Psychotherapist, private practice | $ 146 | Consultation |
| Psychotherapist, primary care | $ 301 | Consultation |
| Psychotherapist, psychiatry | $ 477 | Consultation |
| Counselor | $ 287 | Consultation |
| Physiotherapist | $ 107 | Consultation |
| Midwife | $ 199 | Consultation |
| Alternative care | $ 87 | Consultation |
| Home care | $ 58 | Hour |
| Self-help group | $ 13 | Hour |
| Note. All costs are in 2014-06-01 US dollar, converted from the Swedish krona. Most estimates are based on official public listings in the publicly funded Swedish health care system. | | |

### Supplemental results

#### Detailed cost breakdown

In the tables below, costs from various sources in the TIC-P are shown separately. Costs for providing therapy (i.e., therapist time) are not included. Costs for health care visits are included if they were unrelated to the clinical trial, since all visits in the research study were free of charge for participants.

| Table A.2. TIC-P costs at pre-treatment | | |
| --- | --- | --- |
| Pre-treatment |  |  |
|  | BDD-NET (n = 47) | Supportive psychotherapy (n = 47) |
| Direct medical costs | 1261 (1943), 389 | 1517 (2122), 331 |
| *Healthcare visits* | 1162 (1893), 304 | 1473 (2110), 321 |
| *Medication* | 99 (311), 15 | 43 (57), 9 |
| Direct non-medical costs | 6 (38), 0 | 0 (0), 0 |
| Indirect costs | 3396 (5222), 610 | 2838 (4821), 689 |
| *Unemployment* | 1340 (3540), 0 | 893 (2960), 0 |
| *Sick leave* | 1000 (3367), 0 | 608 (2284), 0 |
| *Work cutback* | 663 (2247), 0 | 808 (2231), 0 |
| *Domestic* | 393 (795), 112 | 528 (1167), 38 |
| **Gross total costs** | **4662 (6249), 2261** | **4354 (5821), 1788** |
| *Note.* Mean (SD), median. All costs are in 2014-06-01 US Dollar and extrapolated to a three-month period. | | |

| Table A.3. TIC-P costs at post-treatment | | |
| --- | --- | --- |
| Post-treatment |  |  |
|  | BDD-NET (n = 45) | Supportive psychotherapy (n = 46) |
| Direct medical costs | 1268 (2661), 2 | 1023 (1766), 115 |
| *Healthcare visits* | 1194 (2619), 0 | 969 (1769), 0 |
| *Medication* | 74 (234), 0 | 54 (202), 7 |
| Direct non-medical costs | 47 (188), 0 | 34 (171), 0 |
| Indirect costs | 3404 (4848), 658 | 2619 (4234), 712 |
| *Unemployment* | 1866 (4057), 0 | 228 (1547), 0 |
| *Sick leave* | 772 (2775), 0 | 869 (2434), 0 |
| *Work cutback* | 455 (1296), 0 | 1131 (2699), 234 |
| *Domestic* | 311 (577), 37 | 390 (731), 65 |
| **Gross total costs** | **4719 (6441), 1233** | **3677 (4781), 1647** |
| *Note.* Mean (SD), median. All costs are in 2014-06-01 US Dollar and extrapolated to a three-month period. | | |

| Table A.4. TIC-P costs at follow-up | | |
| --- | --- | --- |
| Follow-up |  |  |
|  | BDD-NET (n = 36) | Supportive psychotherapy (n = 43) |
| Direct medical costs | 1396 (2208), 588 | 714 (1345), 44 |
| *Healthcare visits* | 1296 (2161), 0 | 679 (1338), 0 |
| *Medication* | 101 (351), 7 | 35 (67), 4 |
| Direct non-medical costs | 0 (0), 0 | 0 (0), 0 |
| Indirect costs | 2941 (4480), 229 | 2767 (4389), 446 |
| *Unemployment* | 1749 (3966), 0 | 1220 (3404), 0 |
| *Sick leave* | 580 (2285), 0 | 319 (1250), 0 |
| *Work cutback* | 372 (1240), 0 | 947 (2208), 0 |
| *Domestic* | 240 (653), 2 | 282 (580), 8 |
| **Gross total costs** | **4337 (5159), 2479** | **3481 (4791), 1164** |
| *Note.* Mean (SD), median. All costs are in 2014-06-01 US Dollar and extrapolated to a three-month period. | | |

#### Cost-effectiveness planes

The supplemental cost-effectiveness planes show costs versus effect at post-treatment and follow-up from all three perspectives. In the societal perspective, all direct and indirect medical and non-medical costs are included; in the direct medical perspective, only direct medical costs are included (i.e., treatment costs, health care visits and medications); in the health organisational perspective, only the cost of providing treatment is included.

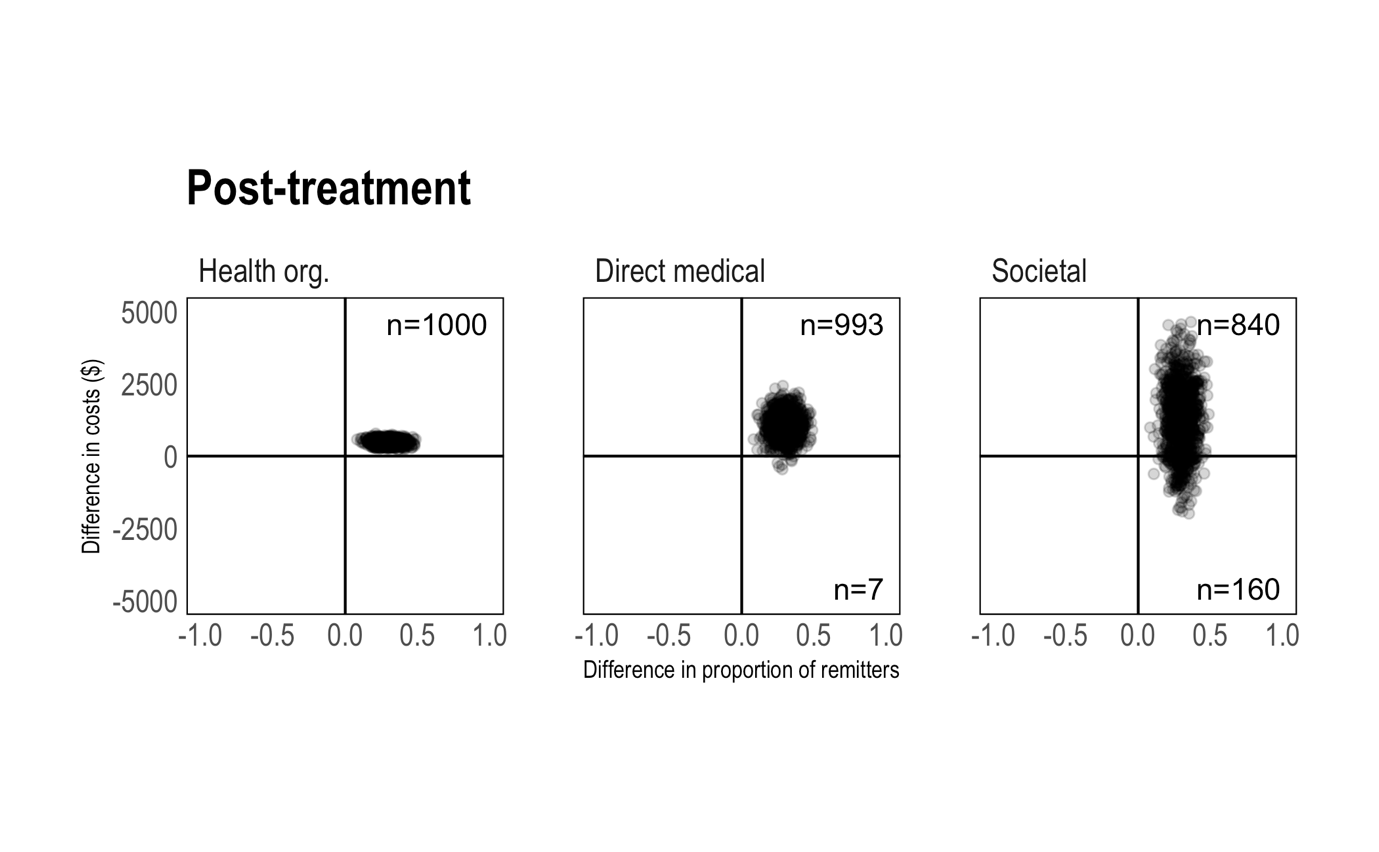

Figure A.1. Cost-effectiveness of BDD-NET versus supportive psychotherapy from pre- to post-treatment. Effect based on proportion of patients in remission after treatment.
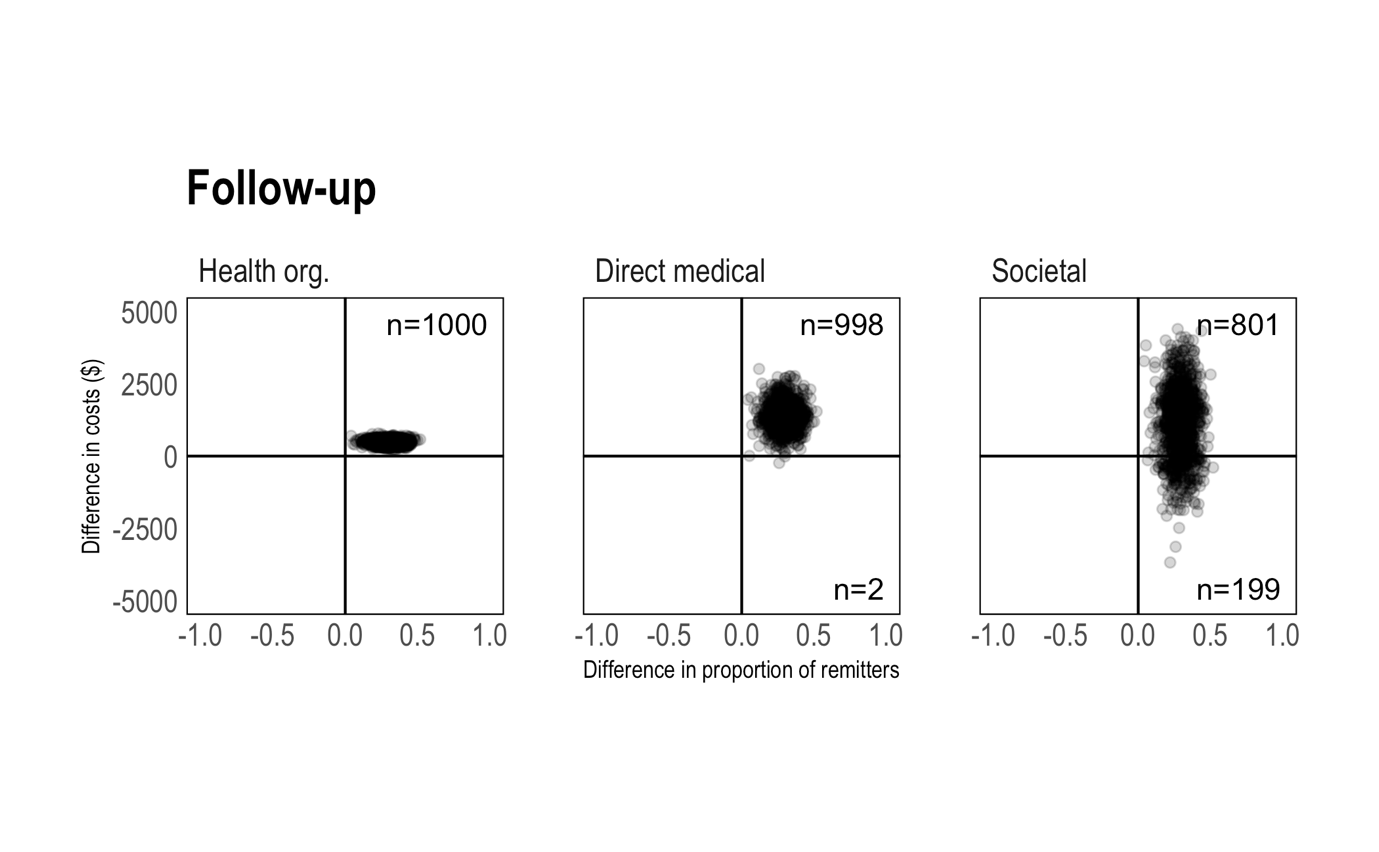
 Figure A.2. Cost-effectiveness of BDD-NET versus supportive psychotherapy from pre-treatment to follow-up. Effect based on proportion of patients in remission at follow-up.

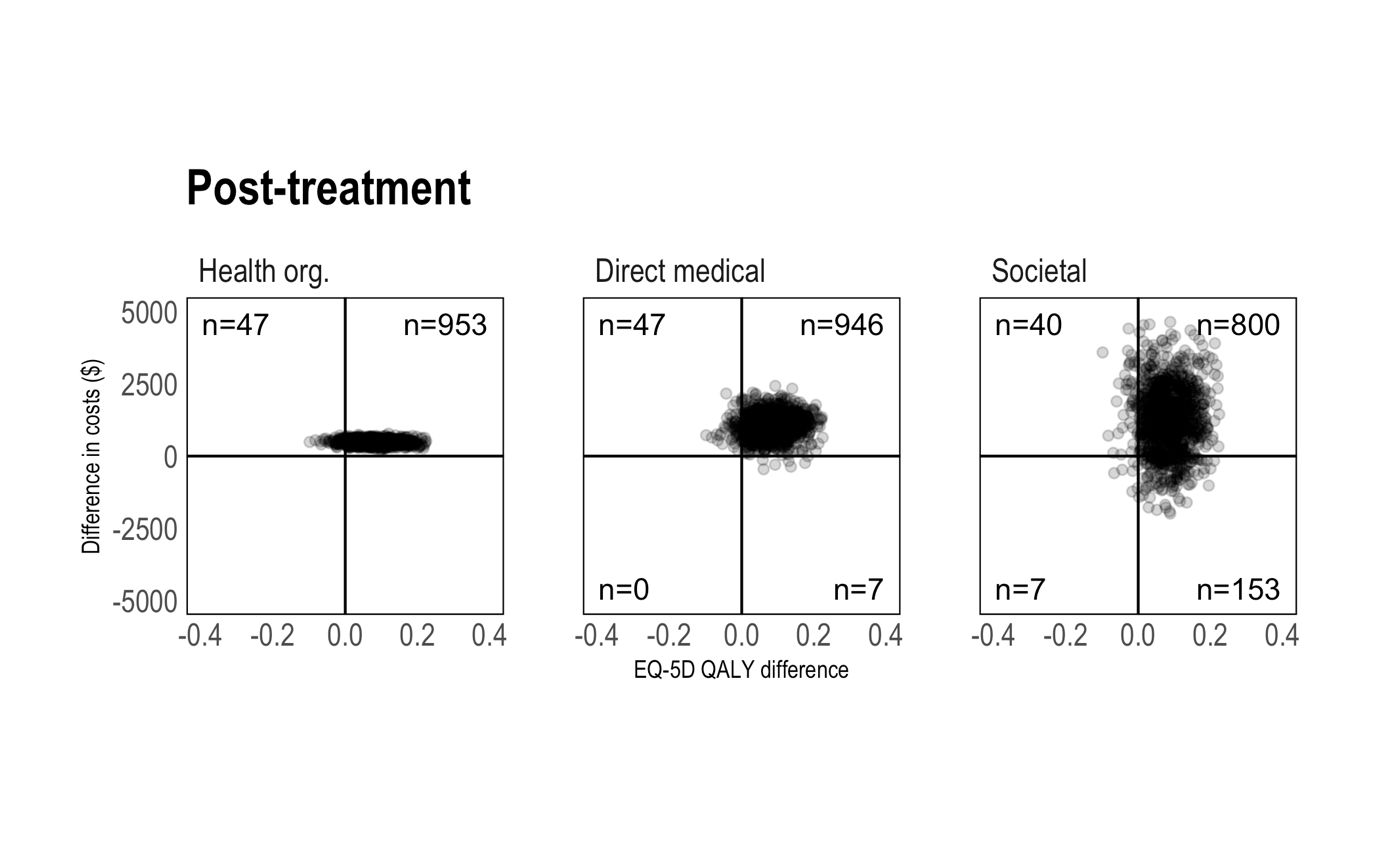

Figure A.3. Cost-effectiveness of BDD-NET versus supportive psychotherapy from pre- to post-treatment. Effect based on quality-adjusted life year (QALY) change from pre- to post-treatment.

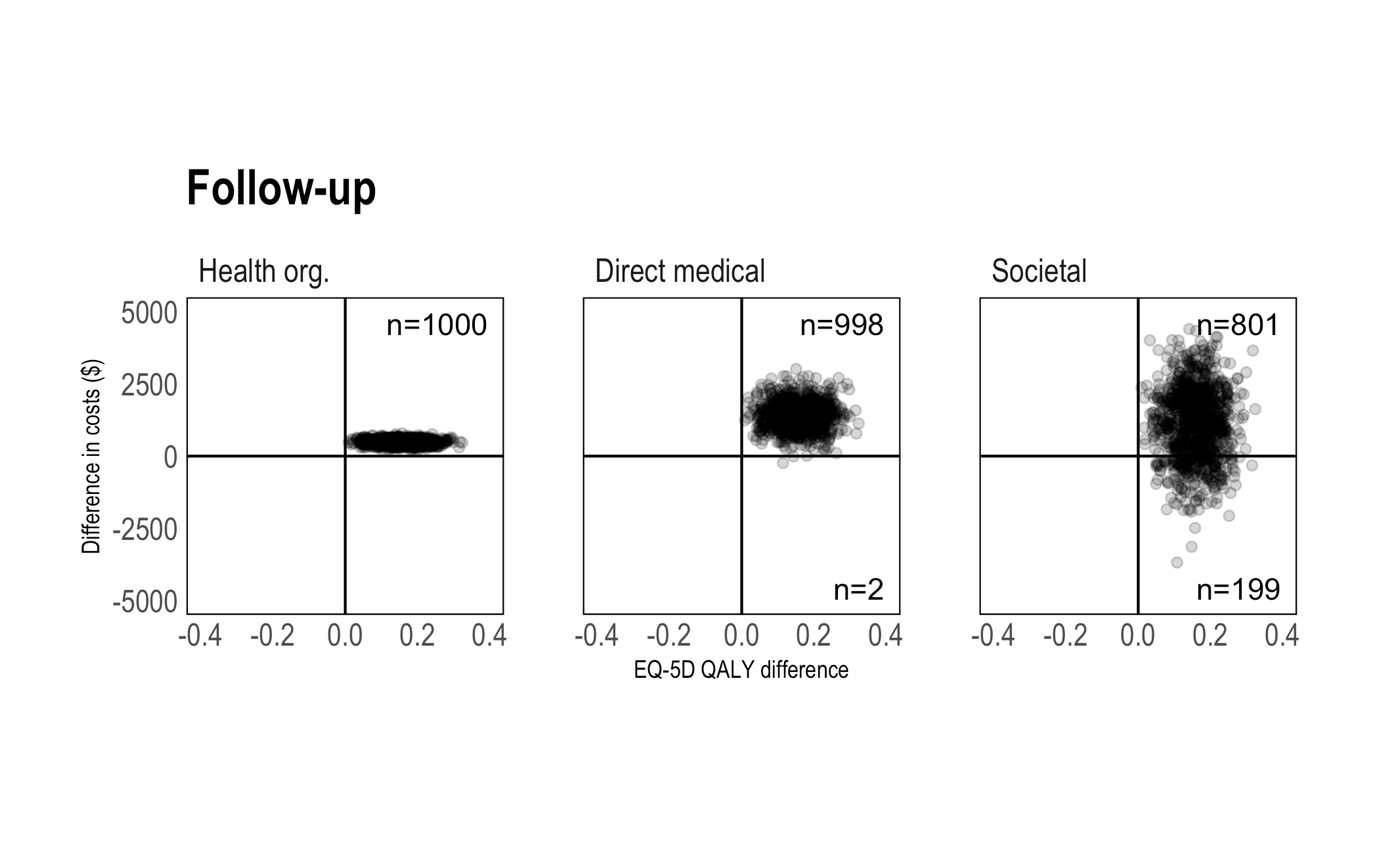

Figure A.4. Cost-effectiveness of BDD-NET versus supportive psychotherapy from pre- to follow-up. Effect based on quality-adjusted life year (QALY) change from pre-treatment to follow-up.

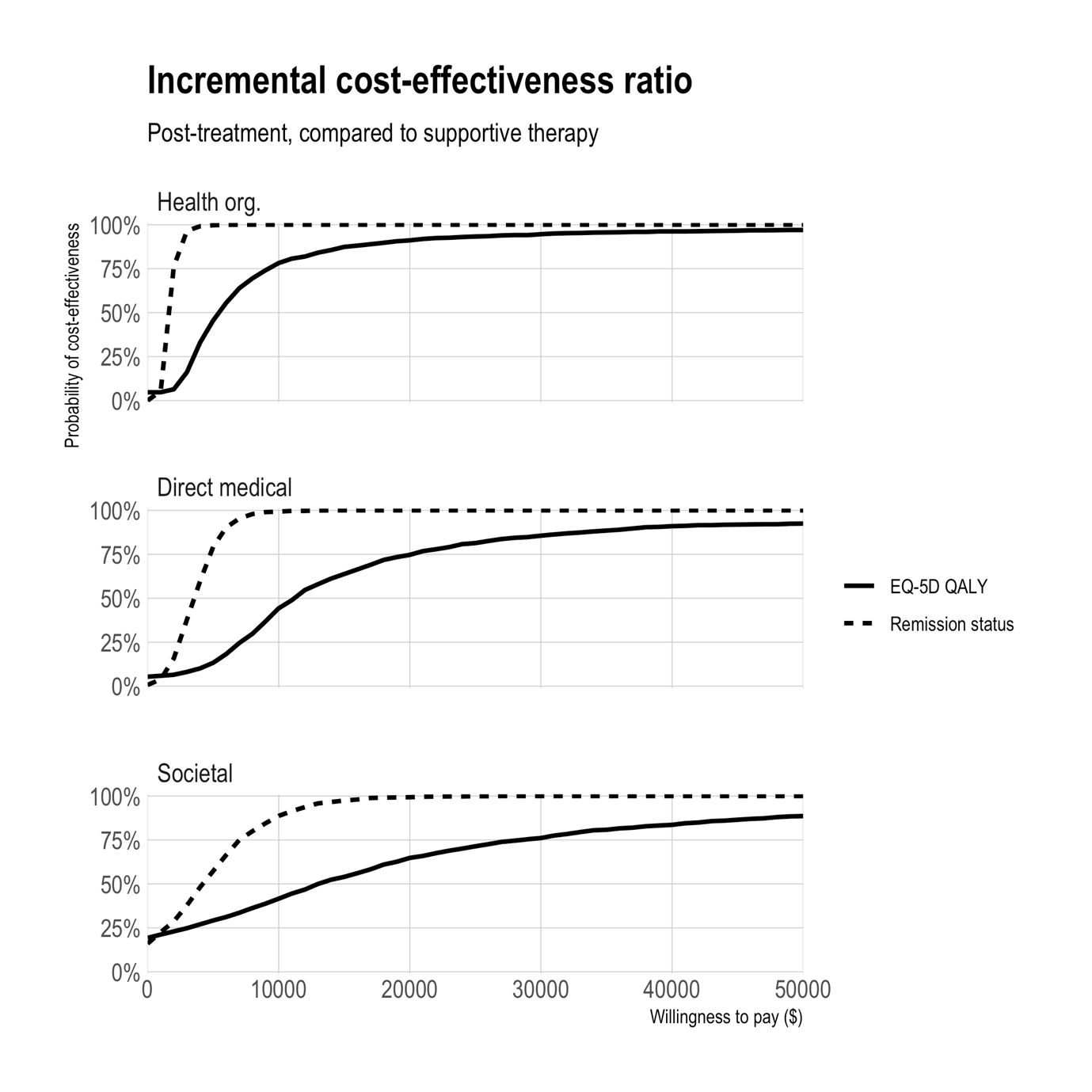

Figure A.5. Acceptability curves at post-treatment for quality-adjusted life years and remission status, with costs from different perspectives.
